# Dual Factors in Chemotherapy Dose Reductions: Bridging Causal Insights with Clinical Correlates

**DOI:** 10.1101/2025.04.23.25326285

**Authors:** Hakan Şat Bozcuk, Mustafa Yıldız, Mehmet Artaç, Hasan Mutlu, Hasan Şenol Coşkun

## Abstract

Chemotherapy dose reductions are associated with adverse clinical outcomes in cancer patients, yet the underlying causal factors remain incompletely defined. We investigated causal determinants of chemotherapy dose reductions across three cancer-type categories (breast, colorectal, and lung/other). We compared these causal determinants with purely correlative factors identified by multivariate Logistic Regression analysis. For causal analysis, we employed the Independent Component Analysis Linear Non-Gaussian Acyclic Model (ICALiNGAM) and all causal analyses were conducted in Python within the Google Colab environment. Among 3,439 chemotherapy cycles, 322 (9%) involved dose reductions. We revealed factors demonstrating both causal and correlative significance (“dual factors”): a prior history of febrile neutropenia, cancer type, a prior history of febrile neutropenia, inpatient treatment and pre-cycle decision to use G-CSF prophylaxis. Our findings reveal distinct causal and correlative factors driving chemotherapy dose reductions, offering clearer targets for intervention to preserve optimal dose intensity.

## INTRODUCTION

Chemotherapy remains a cornerstone of cancer treatment, with dose intensity playing a pivotal role in determining clinical outcomes (1,2). Nevertheless, despite advancements in supportive care, a range of clinical and treatment-related factors often necessitate dose reductions, potentially compromising both efficacy and patient survival (3). Historically, numerous studies have shed light on factors correlated with chemotherapy dose reductions—most notably, advanced age, presence of comorbidities, and extensive prior treatment history (4–6). Although these associations have undoubtedly contributed to our understanding, they have often been confined to correlation, leaving open the critical question of causation (7).

A key principle that has gained increasing attention in medical research is that correlation does not imply causation. Indeed, correlational findings indicate that variables move together, but they provide little insight into whether one variable directly influences another or whether both variables are influenced by an unobserved confounder. To address this gap, methods such as randomized controlled trials (RCTs) are commonly used to rigorously establish causal relationships; however, RCTs may be impractical or unethical in certain clinical contexts. Consequently, advanced statistical and computational techniques, including causal inference modeling, have emerged to bridge this gap (8). In particular, these methods, which leverage non-Gaussian data distributions and structural assumptions, offer the potential to uncover both the directionality and the magnitude of effects among intertwined clinical variables (9).

Meanwhile, previous research has implicated a variety of patient- and treatment-related factors in chemotherapy dose reductions. For instance, patients with higher age or compromised bone marrow reserves and comorbid conditions—such as renal or hepatic dysfunction—are more likely to experience severe hematologic toxicities, ultimately leading to dose modifications (10). Additionally, treatment-related variables, such as multi-agent regimens and more intensive dosing schedules, can predispose patients to higher toxicities, further necessitating dose adjustments (11). Notably, in some cancer types, a prior history of neutropenia or febrile neutropenia has been consistently associated with a greater likelihood of future reductions, underscoring the role of baseline hematologic status in guiding dose intensity (12). Taken together, these findings highlight the importance of distinguishing between factors that simply correlate with dose modifications and those that actively precipitate them, as interventions targeting the latter may prove most effective in preserving therapeutic benefit.

In this context, we planned to apply the Linear Non-Gaussian Acyclic Model (LiNGAM), a novel approach in causal inference, to delineate the direct and indirect factors driving chemotherapy dose reduction (13, 9). Within this framework, the ICALiNGAM algorithm enables the identification and quantification of causal pathways, making it especially well-suited for complex clinical datasets (14). Moreover, we also aimed to compare causal factors with the correlative factors emerging from our dataset. By pinpointing true causal factors rather than mere correlates, or dual factors bearing both causal and correlative significance, we hope to provide actionable insights that inform clinical strategies for minimizing dose reductions, thereby helping to maintain therapeutic efficacy and optimize patient outcomes.

## MATERIALS AND METHODS

Study Design and Data Sources: We conducted a prospective analysis of chemotherapy treatment data from cancer patients with various diagnoses between 2007 and 2011. A preliminary analysis of this dataset was previously presented at the 2014 ESMO meeting (15). Ethical approval had been obtained for the parental study that focused on development on development of a nomogram for febrile neutropenia and analyzed an overlapping dataset for another endpoint, with a parallel research question (16). Due to the purely observational nature of the study, the requirement for consent was not enforced by the ethics committee. For the current study, we utilized the same database, including all cases with at least one cycle of chemotherapy and complete data for all evaluated features. Unlike the earlier analysis, this study adopts a novel approach by incorporating causal analysis alongside traditional statistical and correlative methods.

The study had been approved by the institutional review board, and all data were anonymized prior to analysis. The predictors evaluated in this study were general factors (cancer type, stage, involved systems number – a measure of number of organ systems involved by cancer, gender, age, ECOG performance status, coronary disease status, Chronic Obstructive Lung Disease (COLD) status, previous radiotherapy receipt, receipt of a different protocol of chemotherapy before), treatment factors (treatment as an inpatient, previous febrile neutropenia episode, drug number in the protocol, baseline febrile neutropenia risk classification for the current chemotherapy protocol according to NCCN, cycle no on current protocol) and laboratory factors (precycle values of LDH, ALT, Creatinine, Lymphocyte count and Albumin level). We made use of AI assisted editing to improve the paper (ChatGPT 4o).

Correlative Statistical Analysis: To complement the causal analysis described below, we also performed univariate and multivariate Logistic Regression (LR) to identify non-causal, correlative associations between predictors and chemotherapy dose reductions. Predictors with a P-value less than 0.10 in the univariate LR analysis were subsequently included in the multivariate LR analysis, which employed a Forward Stepwise algorithm based on the Likelihood Ratio test. Statistical significance in the multivariate analysis was defined as a P-value less than 0.05. All LR analyses were conducted using SPSS version 21.

Causal Analysis Framework: We employed ICALiNGAM to identify causal factors for the occurrence of dose reduction. ICALiNGAM is an advanced causal discovery algorithm that enhances the ability to uncover underlying causal structures within complex datasets. By integrating Independent Component Analysis (ICA) with the LiNGAM framework, ICALiNGAM leverages the strengths of both methodologies to effectively identify and estimate the directionality of causal relationships among variables. ICA is utilized to decompose multivariate data into statistically independent components, which is crucial for isolating the unique contributions of each variable without confounding influences. Building upon this, the LiNGAM approach assumes that the data-generating process is linear and that the underlying variables follow non-Gaussian distributions, allowing for the determination of a unique causal ordering that respects the acyclic nature of causal graphs.

This combination enables ICALiNGAM to not only detect the presence of causal links but also to infer the precise direction of causality, which is essential for applications such as dose reduction strategies in healthcare. Additionally, ICALiNGAM is robust in scenarios where traditional methods may struggle, particularly in the presence of latent confounders and when the data exhibits non-Gaussian characteristics. Its ability to produce a comprehensive adjacency matrix that maps out the causal pathways provides valuable insights for decision-making and intervention planning, making ICALiNGAM a powerful tool to understand and manipulate complex causal systems. All analyses were performed in Python using the Google Colab platform. Direct, indirect, and total effects of causal variables on dose reduction were computed using the adjacency matrix. Variables with the highest total effects were identified as key causal factors. Refer to Figure 1 for types of causal effects.

**Figure 1:**
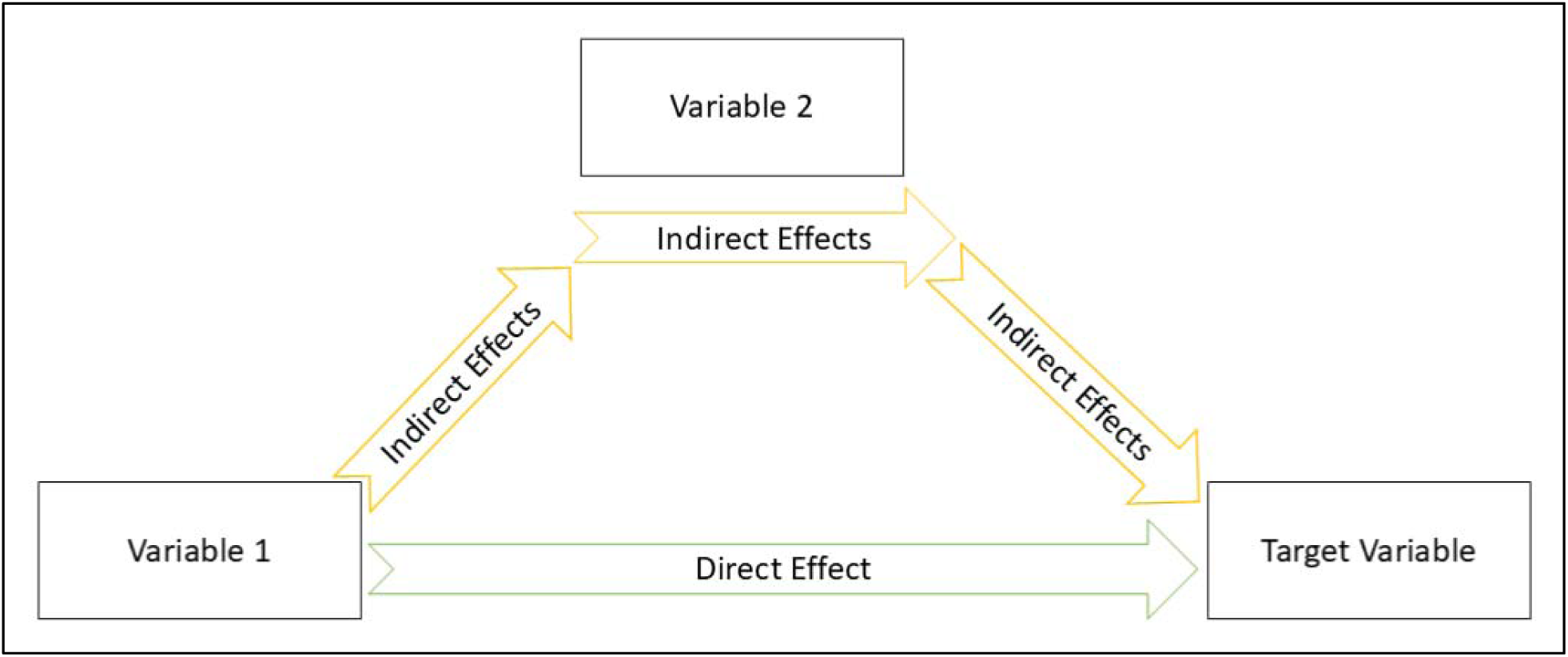
Causal effect types. Direct and indirect effect types on target variable.

## RESULTS

General: Out of 3439 chemotherapy cycles, dose reductions were documented in 322 cycles (9%). Chemotherapy was delivered to female patients in 53% of the cycles, breast cancer was the most frequent diagnosis (39%), and majority of cycles (56%) were delivered to stage 1 to 3 patients. The median age was 55, see Table 1 for details of patient demographics with and without chemotherapy dose reduction.

**Table 1:** Demographic data with respect to chemotherapy dose reduction status.

| Features | All cases |  |  | Cases without dose reduction |  |  | Cases with dose reduction |  |  |
| --- | --- | --- | --- | --- | --- | --- | --- | --- | --- |
|  | n | % | Median | n | % | Median | n | % | Median |
| Chemotherapy cycles | 3439 | 100 |  | 3117 | 100 (91%) |  | 322 | 100 (9%) |  |
| <u>General</u> |  |  |  |  |  |  |  |  |  |
| Cancer type |  |  |  |  |  |  |  |  |  |
| Breast cancer | 1315 | 38 |  | 1226 | 39 |  | 89 | 28 |  |
| Lung cancer | 890 | 26 |  | 803 | 26 |  | 87 | 27 |  |
| Colorectal cancer | 1159 | 34 |  | 1034 | 33 |  | 125 | 39 |  |
| Other cancers | 75 | 2 |  | 54 | 2 |  | 21 | 6 |  |
| Stage |  |  |  |  |  |  |  |  |  |
| 1 to 3 | 1927 | 56 |  | 1804 | 58 |  | 123 | 38 |  |
| 4 | 1512 | 44 |  | 1313 | 42 |  | 199 | 62 |  |
| Involved systems number* |  |  | 0 |  |  | 0 |  |  | 1 |
| Gender |  |  |  |  |  |  |  |  |  |
| Female | 1811 | 53 |  | 1443 | 46 |  | 185 | 58 |  |
| Male | 1628 | 47 |  | 1674 | 54 |  | 137 | 42 |  |
| Age |  |  | 55 |  |  | 55 |  |  | 58 |
| ECOG performance status |  |  | 1 |  |  | 1 |  |  | 1 |
| Coronary disease |  |  |  |  |  |  |  |  |  |
| No | 3133 | 91 |  | 2838 | 91 |  | 295 | 92 |  |
| Yes | 306 | 9 |  | 279 | 9 |  | 27 | 8 |  |
| Chronic obstructive Lung disease (COLD) |  |  |  |  |  |  |  |  |  |
| No | 3252 | 95 |  | 2944 | 94 |  | 308 | 96 |  |
| Yes | 187 | 5 |  | 173 | 6 |  | 14 | 4 |  |
| Radiotherapy |  |  |  |  |  |  |  |  |  |
| Did not receive before | 3372 | 98 |  | 3061 | 98 |  | 311 | 97 |  |
| Received before | 67 | 2 |  | 56 | 2 |  | 11 | 3 |  |
| Previous chemotherapy |  |  |  |  |  |  |  |  |  |
| No | 1922 | 56 |  | 1782 | 57 |  | 140 | 44 |  |
| Yes | 1517 | 44 |  | 1335 | 43 |  | 182 | 56 |  |
| <u>Treatment</u> |  |  |  |  |  |  |  |  |  |
| Treatment as an inpatient |  |  |  |  |  |  |  |  |  |
| No | 3240 | 94 |  | 2923 | 94 |  | 317 | 98 |  |
| Yes | 199 | 6 |  | 194 | 6 |  | 5 | 2 |  |
| Previous febrile neutropenia |  |  |  |  |  |  |  |  |  |
| No | 3211 | 93 |  | 2950 | 95 |  | 261 | 81 |  |
| Yes | 228 | 7 |  | 167 | 5 |  | 61 | 19 |  |
| Drug number** |  |  |  |  |  |  |  |  |  |
| 1 | 336 | 10 |  | 313 | 10 |  | 23 | 7 |  |
| 2 | 1644 | 48 |  | 1482 | 48 |  | 162 | 50 |  |
| 3 | 1076 | 31 |  | 986 | 31 |  | 90 | 28 |  |
| 4 | 332 | 11 |  | 335 | 11 |  | 47 | 15 |  |
| 5 | 1 | 0 |  | 1 | 0 |  | 0 | 0 |  |
| Regimen risk*** |  |  |  |  |  |  |  |  |  |
| 1 | 938 | 27 |  | 840 | 27 |  | 98 | 30 |  |
| 2 | 2157 | 63 |  | 1976 | 63 |  | 181 | 57 |  |
| 3 | 344 | 10 |  | 301 | 10 |  | 43 | 13 |  |
| Cycle no on current protocol |  |  | 3 |  |  | 3 |  |  | 4 |
| <u>Laboratory</u> |  |  |  |  |  |  |  |  |  |
| LDH (IU/ml) |  |  | 343 |  |  | 337 |  |  | 393 |
| ALT (IU/ml) |  |  | 18 |  |  | 17 |  |  | 19 |
| Creatinine (mg/dl) |  |  | 0.71 |  |  | 0.71 |  |  | 0.74 |
| Lymphocyte count (x1000/mm3) |  |  | 1.7 |  |  | 1.74 |  |  | 1.4 |
| Albumin (mg/dl) |  |  | 4.2 |  |  | 4.2 |  |  | 4.2 |

Correlative analysis: To examine the association between various factors and the likelihood of chemotherapy dose reductions, predictors with a P-value of less than 0.10 in univariate analyses were further assessed using multivariate Logistic Regression (LR) models, stratified by cancer type. Several statistically significant factors were identified:

- Breast Cancer: Significant predictors included stage 4 disease (Wald = 13.93, P < 0.001), pre-cycle decision to use G-CSF prophylaxis (Wald = 6.44, P = 0.011), previous febrile neutropenia (Wald = 58.02, P < 0.001), LDH levels (Wald = 3.93, P = 0.047), cycle number in the current protocol (Wald = 36.25, P < 0.001), and ALT levels (Wald = 4.29, P = 0.038).
- Lung or Other Cancers: Significant factors included cancer type (Wald = 12.60, P < 0.001), pre-cycle decision to use G-CSF prophylaxis (Wald = 16.09, P < 0.001), previous febrile neutropenia (Wald = 17.14, P < 0.001), ECOG performance status (Wald = 4.89, P = 0.027), cycle number in the current protocol (Wald = 36.25, P < 0.001), and the number of drugs in the treatment protocol (Wald = 5.72, P = 0.017).
- Colorectal Cancer: Significant factors included inpatient treatment (Wald = 12.30, P < 0.001), prior receipt of chemotherapy (Wald = 7.19, P = 0.007), pre-cycle decision to use G-CSF prophylaxis (Wald = 29.83, P < 0.001), albumin levels (Wald = 10.66, P = 0.001), lymphocyte count (Wald = 9.11, P = 0.003), and cycle number in the current protocol (Wald = 14.55, P < 0.001). Further details of the LR analysis are provided in Table 2.

**Table 2:**
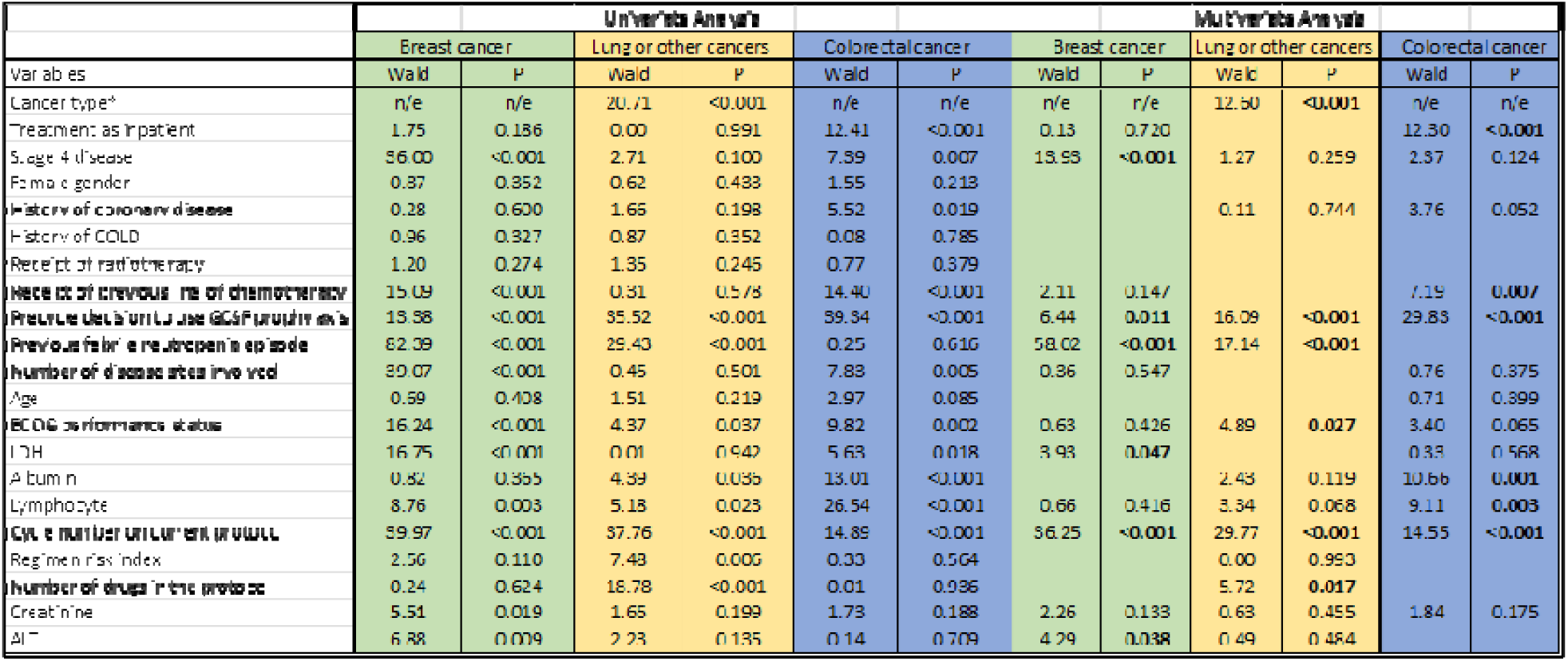
Logistic regression analysis of dose reduction according to cancer type. Univariate and multivariate analysis of correlates of chemotherapy dose reduction in 3 categories of cancer type are presented.

Causal analysis: For the causal analysis of chemotherapy dose reductions in breast cancer using the ICALiNGAM model, significant total effects (te) with a magnitude of ≥ 0.10 were identified for treatment as an inpatient (te = 0.18), previous febrile neutropenia episodes (te = 0.32), and creatinine levels (te = -0.11). In the lung or other cancer subgroup, significant causal factors included cancer type (te = -0.22) and previous febrile neutropenia episodes (te = 0.20). For colorectal cancer, key causal factors contributing to dose reductions included treatment as an inpatient (te = -0.11), receipt of radiotherapy (te = 0.14), and pre-cycle decisions to use G-CSF prophylaxis (te = 0.21).

Detailed information on direct, indirect, and total causal effects is provided in Table 3. Additionally, Figures 2, 3, and 4 illustrate the adjacency matrices and causal networks for the breast, lung or other, and colorectal cancer subgroups, respectively.

**Figure 2:**
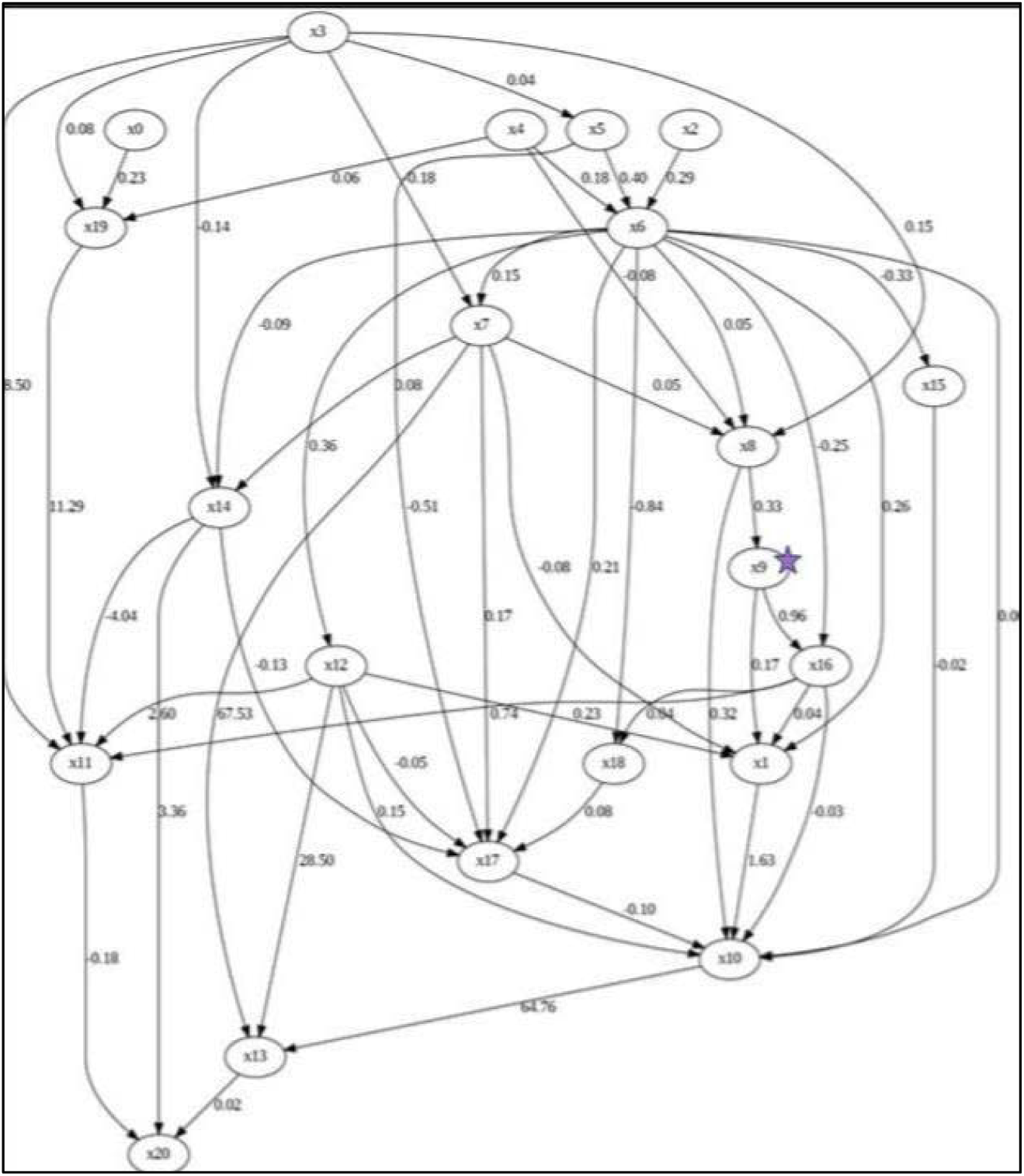
Causal graph for dose reduction in breast cancer. Factors with causal effect on dose reduction in breast cancer. Star sign indicates the variable for dose reduction. X0; Treatment as inpatient, x1; stage 4, x2; female gender, x3; coronary disease, x4; chronic obstructive lung disease, x5; Radiotherapy, x6; previous chemotherapy, x7; Colony Stimulating Factor usage, x8; previous febrile neutropenia, x9; dose reduction, x10; number of Involved systems, x11; age, x12; ECOG performance status, x13; LDH, x14; albumin, x15; lymphocyte count, x16; Cycle no on current protocol, x17; regimen risk, x18; drug number, x19; creatinine, x20; ALT

**Figure 3:**
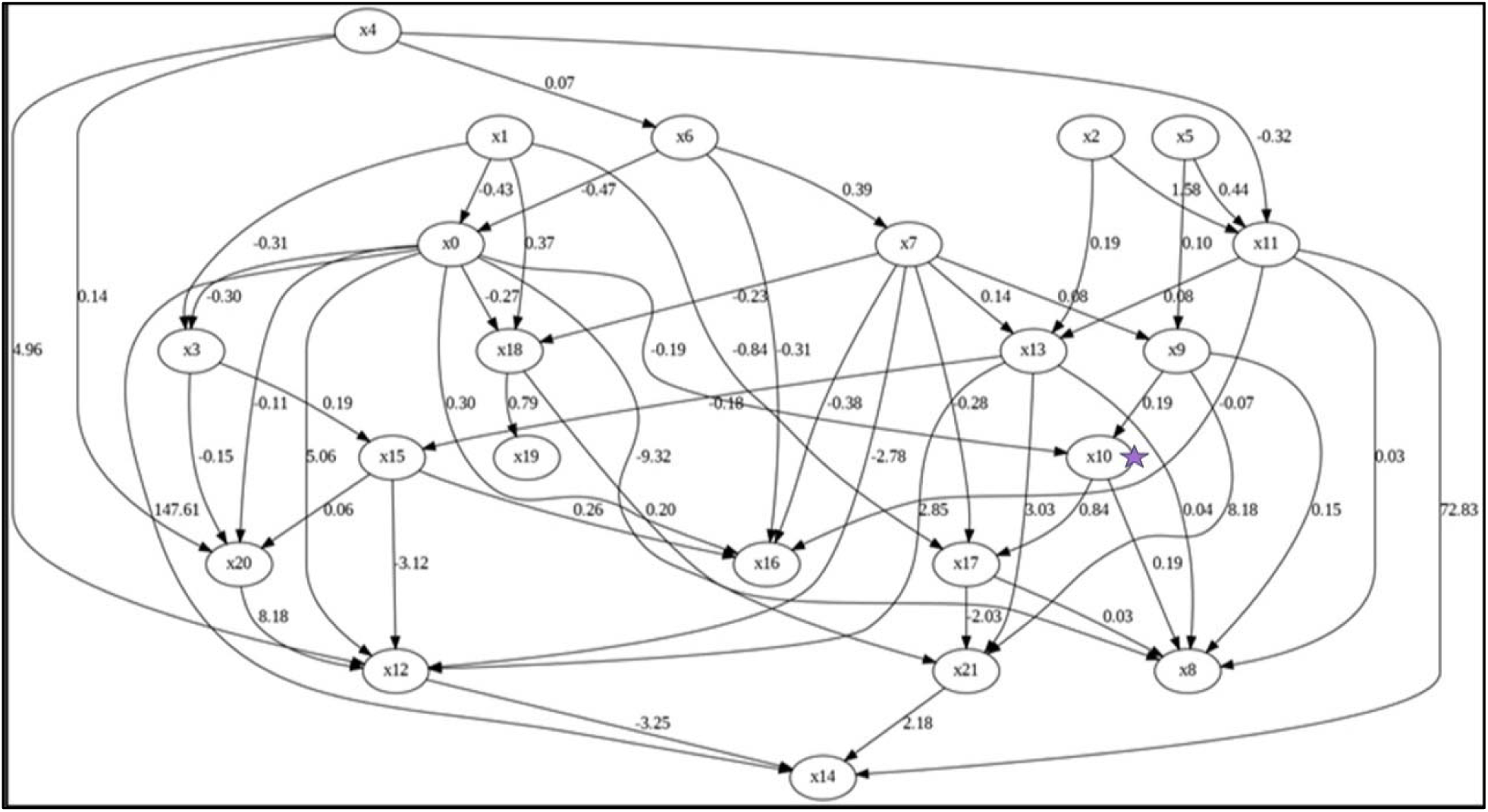
Causal graph for dose reduction in lung or other cancers. Factors with causal effect on dose reduction in lung or other cancers. Star sign indicates the variable for dose reduction. X0; type of cancer (lung cancer versus other cancers), X1; treatment as inpatient, x2; stage 4, x3; female gender, x4; coronary disease, x5; chronic obstructive lung disease, x6; Radiotherapy, x7; previous chemotherapy, x8; Colony Stimulating Factor usage, x9; previous febrile neutropenia, x10; dose reduction, x11; number of Involved systems, x12; age, x13; ECOG performance status, x14; LDH, x15; albumin, x16; lymphocyte count, x17; Cycle no on current protocol, x18; regimen risk, x19; drug number, x20; creatinine, x21; ALT

**Figure 4:**
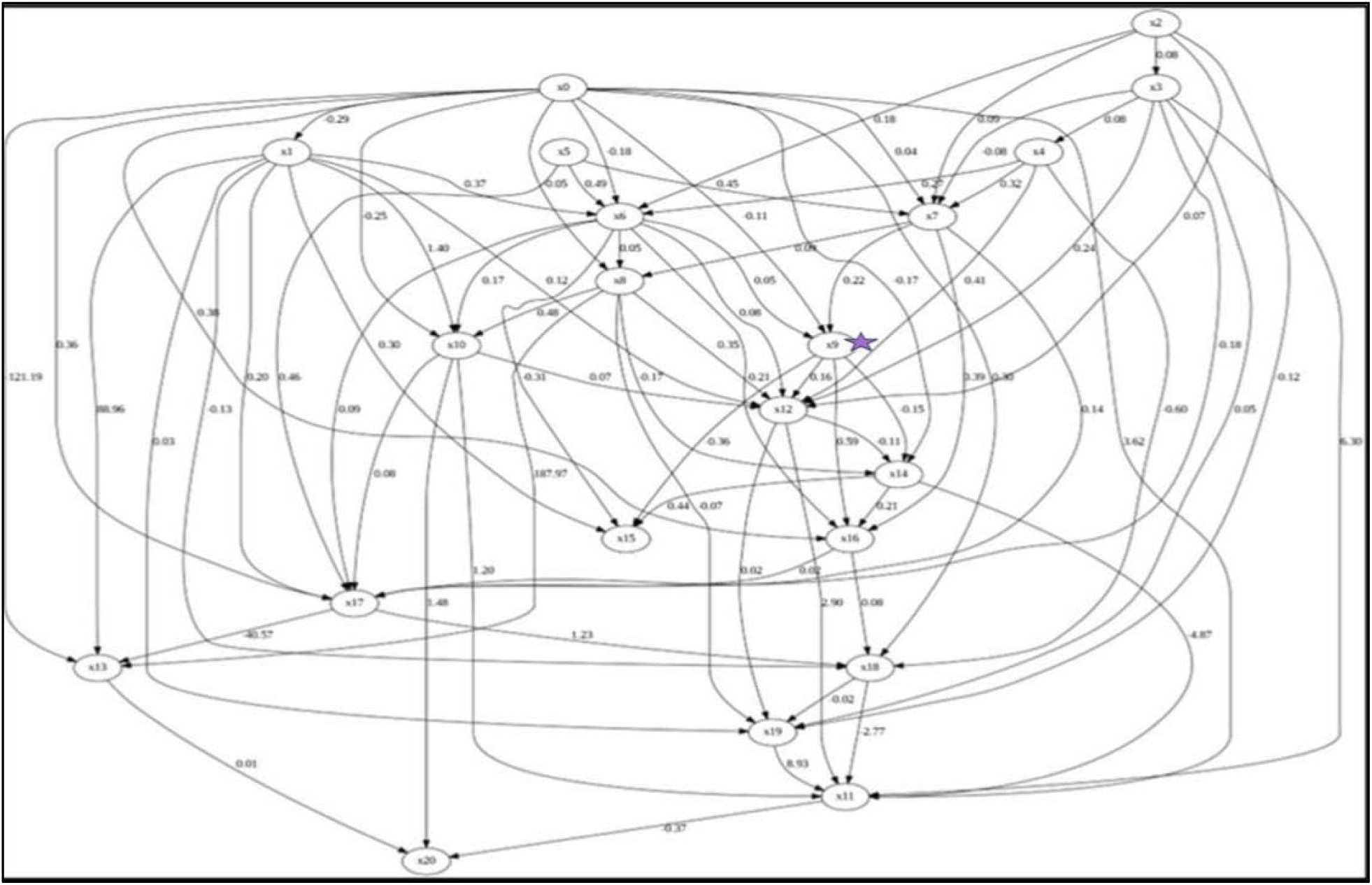
Causal graph for dose reduction in colorectal cancer. Factors with causal effect on dose reduction in colorectal cancer. Star sign indicates the variable for dose reduction. X0; Treatment as inpatient, x1; stage 4, x2; female gender, x3; coronary disease, x4; chronic obstructive lung disease, x5; Radiotherapy, x6; previous chemotherapy, x7; Colony Stimulating Factor usage, x8; previous febrile neutropenia, x9; dose reduction, x10; number of Involved systems, x11; age, x12; ECOG performance status, x13; LDH, x14; albumin, x15; lymphocyte count, x16; Cycle no on current protocol, x17; regimen risk, x18; drug number, x19; creatinine, x20; ALT

**Table 3:**
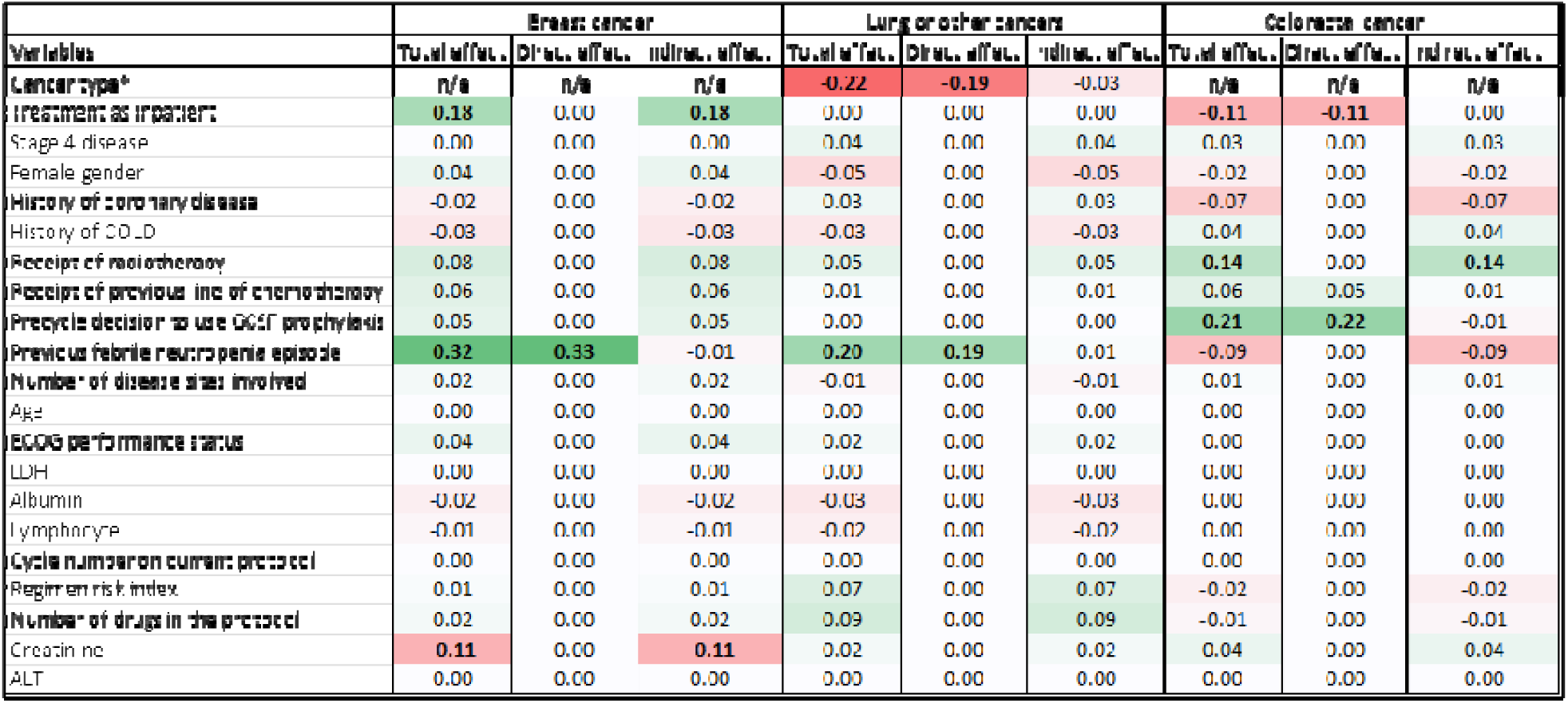
Causal factors on chemotherapy dose reduction according to cancer type. Direct, indirect and total effect on chemotherapy dose reduction of various factors in different cancer types.

Dual important factors: When evaluating both correlative and causal factors for chemotherapy dose reductions, certain factors were identified as having both correlative and causal significance. For the breast cancer subgroup, the key factor was a previous febrile neutropenia episode. In the lung and other cancer subgroup, both previous febrile neutropenia episodes and cancer type were significant. In the colorectal cancer subgroup, inpatient treatment status and pre-cycle decisions to use G-CSF prophylaxis were notable dual factors. These findings are summarized in Table 4, with Figure 5 illustrating their relationship to chemotherapy dose reductions.

**Figure 5:**
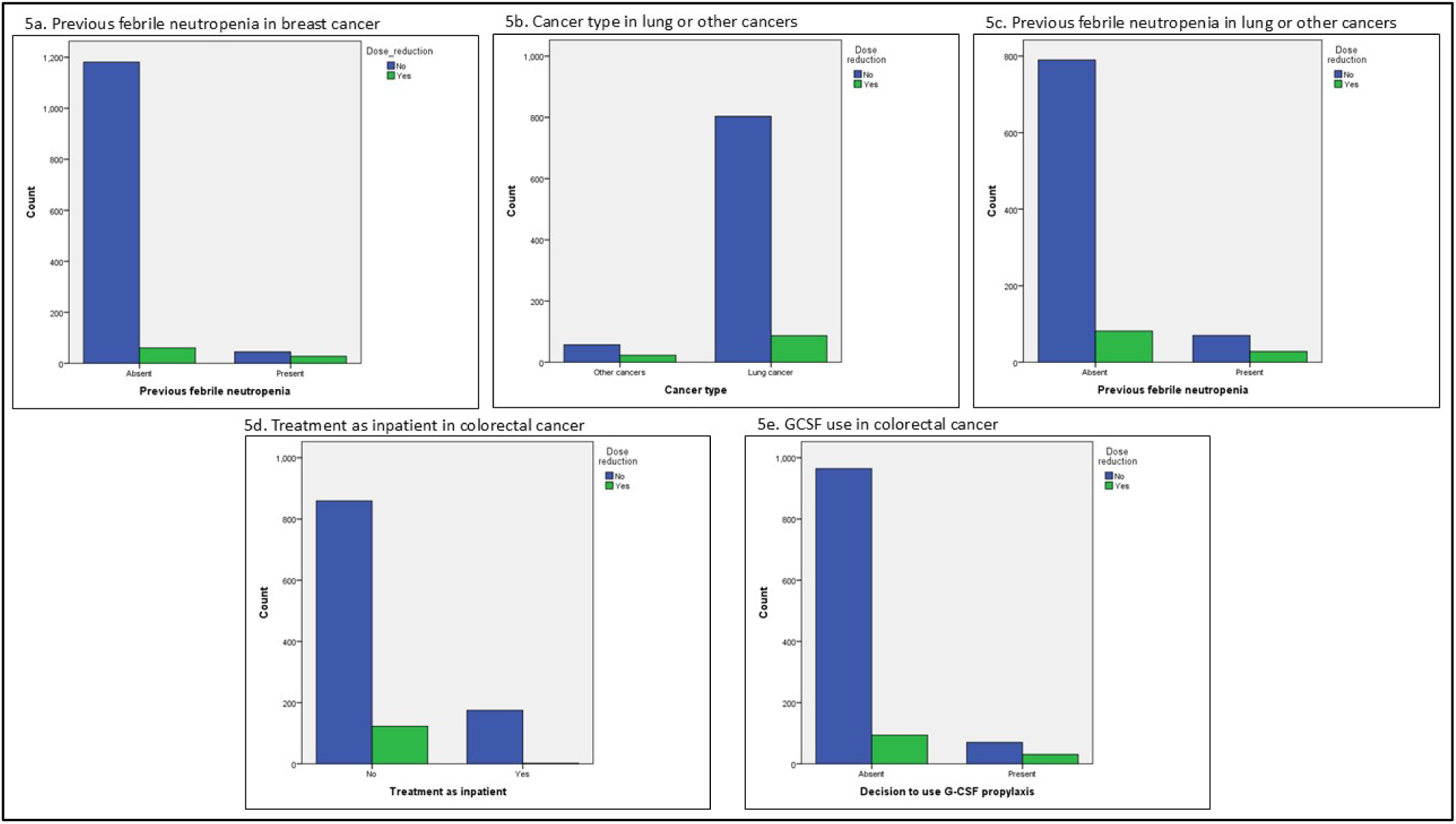
Significant dual correlative and causal factors for chemotherapy dose reduction. 5a. Previous febrile neutropenia in breast cancer, 5b. Cancer type in lung or other cancers, 5c. Previous febrile neutropenia in lung or other cancers, 5d. Treatment as inpatient in colorectal cancer, 5e. GCSF use in colorectal cancer.

**Table 4:** Association of correlative and causal analysis. Factors that possess both correlative and causal significance on chemotherapy dose reduction in different cancer types. Blue color represents dual significance.

| Variables | Breast cancer |  | Lung or other cancers |  | Colorectal cancer |  |
| --- | --- | --- | --- | --- | --- | --- |
|  | Correlative | Causal | Correlative | Causal | Correlative | Causal |
| Cancer type* | n/e | n/e | significant | significant | n/e | n/e |
| Treatment as inpatient |  | significant |  |  | significant | significant |
| Stage 4 disease | significant |  |  |  |  |  |
| Female gender |  |  |  |  |  |  |
| History of coronary disease |  |  |  |  |  |  |
| History of COLD |  |  |  |  |  |  |
| Receipt of radiotherapy |  |  |  |  |  | significant |
| Receipt of previous line of chemotherapy |  |  |  |  | significant |  |
| Pre-cycle decision to use G-CSF prophylaxis | significant |  | significant |  | significant | significant |
| Previous febrile neutropenia episode | significant | significant | significant | significant |  |  |
| Number of disease sites involved |  |  |  |  |  |  |
| Age |  |  |  |  |  |  |
| ECOG performance status |  |  | significant |  |  |  |
| LDH | significant |  |  |  |  |  |
| Albumin |  |  |  |  | significant |  |
| Lymphocyte |  |  |  |  | significant |  |
| Cycle number on current protocol | significant |  | significant |  | significant |  |
| Regimen risk Index |  |  |  |  |  |  |
| Number of drugs in the protocol |  |  | significant |  |  |  |
| Creatinine |  | significant |  |  |  |  |
| ALT | significant |  |  |  |  |  |

## DISCUSSION

Our study emphasizes the importance of applying causal inference methods in oncology, identifying in this study factors such as febrile neutropenia history, G-CSF use, and prior radiotherapy as key drivers of chemotherapy dose reductions. These results highlight the critical need to differentiate causal relationships from mere associations to inform the development of targeted and effective interventions (179.

The direct effect of febrile neutropenia history on chemotherapy dose reductions highlights the importance of proactive strategies such as dose adjustments or prophylactic G-CSF use for patients with prior neutropenic events (18). Patients with a history of febrile neutropenia face an elevated risk of recurrent episodes, which can result in chemotherapy delays, dose reductions, and potentially compromised treatment outcomes. Proactive management approaches, including dose modifications and the prophylactic administration of granulocyte-colony stimulating factors (G-CSF), have proven effective in reducing the incidence and severity of febrile neutropenia among high-risk patients.

Prophylactic G-CSF not only lowers the risk of hospitalization and infection-related complications but also helps sustain chemotherapy dose intensity, thereby enhancing treatment adherence and efficacy (19,20). Furthermore, individualized risk assessments that account for a history of febrile neutropenia enable tailored interventions to optimize clinical outcomes while mitigating toxicity (16). Integrating these preventive strategies into treatment plans for patients with febrile neutropenia history is a vital aspect of supportive oncology care.

Cancer type emerged as a key dual factor in our analysis, demonstrating both correlative and causal associations with chemotherapy dose reductions. Different cancer types influence chemotherapy tolerance and dose intensity requirements due to variations in tumor biology, treatment regimens, and patient-specific factors. Research indicates that patients with certain cancers such as certain breast cancer subtypes, ovarian cancer and colorectal cancer may be faced with increased mortality risk after chemotherapy dose reduction (21,22). These findings underscore the importance of tailoring dose adjustment strategies not only to individual patient characteristics but also to the specific cancer type being treated, ensuring optimal therapeutic outcomes while minimizing adverse effects.

Inpatient treatment status demonstrated both correlative and causal significance in relation to chemotherapy dose reductions. Patients receiving inpatient care often have more advanced disease, comorbidities, or complications requiring hospital-based management, all of which can impact treatment tolerance and necessitate dose modifications (23). From a causal perspective, inpatient status may serve as a proxy for underlying factors such as disease severity, impaired organ function, or reduced performance status, which directly influence chemotherapy pharmacokinetics and toxicity risk. However, in contrast to this general trend, our study found that hospitalized colorectal cancer patients who received chemotherapy were less likely to experience dose reductions compared to outpatients.

Specifically, chemotherapy dose reductions occurred in 12.5% of outpatients versus only 1.1% of inpatients with colorectal cancer. This suggests that inpatients receiving chemotherapy were relatively fit and closely monitored for treatment-related toxicities during and after hospitalization. Indeed, the distribution of ECOG performance scores in our study indicated that inpatients had better functional status (ECOG 0 and 1: 98.9%) compared to outpatients (ECOG 0 and 1: 92%) within the colorectal cancer subgroup. These findings underscore the need for individualized treatment planning for inpatients, incorporating a multidisciplinary approach to optimize dose intensity while mitigating the risk of adverse events.

The precycle decision to use granulocyte colony-stimulating factor (G-CSF) prophylaxis emerged as another factor of dual importance, demonstrating both correlative and causal relevance to chemotherapy dose reduction. Prophylactic G-CSF administration is commonly employed to mitigate the risk of febrile neutropenia, a potentially life-threatening complication of chemotherapy. Correlatively, its use may signify a pre-existing concern about a patient’s ability to tolerate full-dose chemotherapy, based on factors such as advanced age, poor performance status, or comorbidities. Causally, G-CSF can influence hematopoietic recovery, enabling the maintenance of dose intensity in subsequent chemotherapy cycles, even in high-risk patients (19, 24). These findings highlight the integral role of G-CSF prophylaxis in chemotherapy management, serving both as a marker of anticipated dose challenges and as a therapeutic intervention to reduce dose-limiting toxicities.

Compared to traditional statistical methods, the Independent Component Analysis with Linear Non-Gaussian Acyclic Model (ICALiNGAM) offers a robust framework for causal analysis by enabling the identification of both direct and mediated pathways between variables. Unlike correlation-based approaches, ICALiNGAM leverages non-Gaussianity to disentangle complex causal relationships, allowing for a more nuanced understanding of underlying mechanisms (14,25). This feature is particularly advantageous when traditional methods struggle to address latent confounders or when the dataset exhibits intricate dependencies. In our work, ICALiNGAM proved instrumental in uncovering critical causal pathways influencing chemotherapy dose reduction, particularly in disentangling direct effects, such as treatment setting or G-CSF use, from mediated effects. Despite potential limitations in certain datasets, the method’s capacity to uncover actionable causal factors highlights its significance in addressing complex biomedical questions, as demonstrated in our study.

Future research should focus on translating these findings into clinical practice by developing strategies to address causal factors and maintain chemotherapy dose intensity. Additionally, the integration of machine learning with causal inference methods presents exciting opportunities for advancing personalized oncology care.

## Data availability

Our dataset can be assessed at https://github.com/hbozcuk/dose_reduction.

## Ethics approval

Not required due to the pure observative nature of the study.

## Patient consent statement

Not required due to the pure observative nature of the study. Permission to reproduce material from other sources: Not required.

## Clinical trial registration

Not required.

## CRediT CLASSIFICATION

Hakan Şat Bozcuk; Conceptualization (lead), Formal Analysis (lead), Methodology (lead), Software (lead), Writing – Original Draft Preparation (lead), Writing – Review & Editing (equal), Mustafa Yıldız; Data Curation (equal), Writing – Review & Editing (equal), Mehmet Artaç; Data Curation (equal), Writing – Review & Editing (equal), Hasan Mutlu; Data Curation (equal), Writing – Review & Editing (equal), Hasan Şenol Coşkun; Data Curation (equal), Writing – Review & Editing (equal).

## REFERENCES

1. Lyman GH. Impact of chemotherapy dose intensity on cancer patient outcomes. J Natl Compr Canc Netw, 2009; 7(1): 99–108. DOI: 10.6004/jnccn.2009.0009.

2. Loibl S, Skacel T, Nekljudova V, et al. Evaluating the impact of Relative Total Dose Intensity (RTDI) on patients’ short and long-term outcome in taxane- and anthracycline-based chemotherapy of metastatic breast cancer- a pooled analysis. BMC Cancer, 2011; 11: 131.

3. Lyman GH. Chemotherapy Dose Intensity and Quality Cancer Care. Oncology, 2006; 20(14): Suppl_9

4. Seve P, Sawyer M, Hanson J, et al. The influence of comorbidities, age, and performance status on the prognosis and treatment of patients with metastatic carcinomas of unknown primary site: a population-based study. Cancer, 2006;106(9):2058–66. doi: 10.1002/cncr.21833.

5. George M, Smith A, Sabesan S, et al. Physical comorbidities and their relationship with cancer treatment and its outcomes in older adult populations: Systematic review. JMIR Cancer, 2021; 7(4): e26425.

6. Repetto L. Greater risks of chemotherapy toxicity in elderly patients with cancer. The Journal of Supportive Oncology, 2003; 1(4 Suppl 2):18–24.

7. Bhandari P. Correlation vs. Causation | Difference, Designs & Examples. Available at “https://www.scribbr.com/methodology/correlation-vs-causation/”, 2023. Accessed at 1/26/2025.

8. Nogueira AR, Pugnana A, Ruggieri S, et al. Methods and tools for causal discovery and causal inference. Available at “https://wires.onlinelibrary.wiley.com/doi/10.1002/widm.1449”, 2022. Accessed at 1/26/2025.

9. Glymour C, Zhang K, Spirtes P. Review of Causal Discovery Methods Based on Graphical Models. Front Genet 2019;10: Article 524. DOI: 10.3389/fgene.2019.00524.

10. Ian I. Chemotherapy for elderly patients with advanced cancer: is it worth it? Aust Prescr 2000; 23: 80–2.

11. Lustberg MB, Kuderer NM, Desai A, et al. Mitigating long-term and delayed adverse events associated with cancer treatment: implications for survivorship. Nat Rev Clin Oncol, 2023: 5: 1– 16.

12. Pettengell R, Schwenkglenks M, Leonard R, et al. Neutropenia occurrence and predictors of reduced chemotherapy delivery: results from the INC-EU prospective observational European neutropenia study. Support Care Cancer, 2008: 16: 1299–1309.

13. Shimizu S, Inazumi T, Sogawa Y, et al. DirectLiNGAM: A Direct Method for Learning a Linear Non-Gaussian Structural Equation Model. J Mach Learn Res, 2011; 12: 1225–1248.

14. Shimizu S, Hoyer PO, Hyvarinen A, et al. A Linear Non-Gaussian Acyclic Model for Causal Discovery. J Mach Learn Res, 2006; 7: 2003–2030.

15. Bozcuk H, Yıldız M, Ucar S, et al. The correlates of dose reduction in chemotherapy for patients with common cancers. a prospective study. Ann Oncol 2014; 25(4): iv80, 241P. Available at “https://www.annalsofoncology.org/article/S0923-7534(19)51668-4/fulltext”.

16. Bozcuk H, Yıldız M, Artaç M, et al. A prospectively validated nomogram for predicting the risk of chemotherapy-induced febrile neutropenia: a multicenter study. Support Care Cancer, 2015; 23(6): 1759–1767.

17. Altman N, Krzywinski M. Association, correlation and causation. Nat Methods, 2015; 12: 899– 900.

18. Cooper KL, Madan J, Whyte S, et al. Granulocyte colony-stimulating factors for febrile neutropenia prophylaxis following chemotherapy: systematic review and meta-analysis. BMC Cancer, 2011; 11:404. DOI: 10.1186/1471-2407-11-404.

19. Kuderer NM, Dale DC, Crawford J, et al. Impact of primary prophylaxis with granulocyte colony-stimulating factor on febrile neutropenia and mortality in adult cancer patients receiving chemotherapy: a systematic review. J Clin Oncol, 2007; 25(21): 3158–67. DOI: 10.1200/JCO.2006.08.8823.

20. Gascon P, Awada A, Karihtala P, et al. Optimal use of granulocyte colony-stimulating factor prophylaxis to improve survival in cancer patients receiving treatment. Wien Klin Wochenschr, 2023; 136(11-12):362–368. DOI: 10.1007/s00508-023-02300-6.

21. Denduluri N, Lyman GH, Wang Y, et al. Chemotherapy dose intensity and overall survival among patients with advanced breast or ovarian cancer. Clin Breast Cancer, 2018; 18(5): 380–386. DOI: 10.1016/j.clbc.2018.02.003.

22. Abidin MNZ, Omar MS, Islahudin F, et al. The survival impact of palliative chemotherapy dose modifications on metastatic colon cancer. BMC Cancer, 2022; 22: 731.

23. Owens PL, Liang L, Barrett ML, et al. Statistical Brief #303; Comorbidities associated with adult inpatient stays, 2019. Healthcare Cost and Utilization Project (HCUP) Statistical Briefs, 2022. Available at “https://www.ncbi.nlm.nih.gov/books/NBK588380/”. Accessed at 1/26/2025.

24. Cornes P, Gascon P, Chan S, et al. Systematic review and meta-analysis of short-versus long-acting Granulocyte Colony-Stimulating Factors for reduction of chemotherapy-Induced febrile neutropenia. Adv Ther, 2018; 35: 1816–1829.

25. Niu W, Gao Z, Song L, et al. Comprehensive review and empirical evaluation of causal discovery algorithms for numerical data. arXiv 2024; 2407.13054v2 [cs.AI]. Available at “https://arxiv.org/html/2407.13054v2”. Accessed at 1/26/2025.

